# Stellate Ganglion Block to Treat Long COVID-19 Syndrome, A 41 patient Retrospective Cohort Study

**DOI:** 10.1101/2023.08.10.23290338

**Authors:** Lisa Pearson, Alfred Maina, Leah Thompson, Sherri Harden, Abbey Aaroe, Taylor Compratt

## Abstract

Post Covid-19 condition (PCC), long COVID-19 syndrome and post-acute sequelae of SARS-CoV-2 (PASC) all refer to a constellation of symptoms that are unresolved long after the acute phase of the viral infection. The severity of symptoms can vary from mild and tolerable to severe and debilitating.^1,2^ Due to the evolving nature of the SARS-CoV-2 pandemic, treatment protocols for the illness are in a constant state of evolution. The early stage of long COVID-19 syndrome contributes to a dearth of treatment protocols based on empirical evidence, while the absence of a conclusive pathophysiological understanding further complicates the development of such protocols. Current treatment regimens include homeopathic medicine, specialist system focused treatments, infusion therapies, hyperbaric oxygenation, and polypharmacy. The physiological, psychological, and societal impact of long COVID-19 cannot be approached casually and must govern the intensity with which the healthcare community approaches treatment of long COVID-19 syndrome.

In this 41-patient cohort study from a chronic pain management practice, the use of either unilateral or bilateral stellate ganglion block (SGB) was explored to manage symptoms associated with long COVID-19 syndrome. Results indicated that a substantial proportion of patients (86%) experienced a reduction of their symptoms following SGB treatment.

## Introduction

Long COVID-19 encompasses a range of persistent symptoms experienced by individuals following an acute long COVID-19 infection. The Centers for Disease Control and Prevention (CDC) defines long COVID-19 as the presence of symptoms persisting for three or more months after initial viral contraction, with no prior existence of these symptoms.^1^ Data collected by the U.S. Census Bureau and analyzed by the CDC’s National Center for Health Statistics (NCHS) revealed that approximately 7.5% of adults in the United States, or roughly 1 in 13 individuals, are affected by long COVID-19 symptoms.^1^ Research has documented diverse prevalence rates of post-COVID symptoms persisting beyond five weeks post infection, with some studies indicating rates as high as 91% in affected patients.^3,4^

Common manifestations associated with long COVID-19, as outlined by the CDC, include fatigue interfering with daily activities, exacerbation of symptoms following physical or mental exertion, fever, breathing difficulties or shortness of breath, cough, chest pain, palpitations, cognitive impairment, headaches, sleep disturbances, orthostatic dizziness, paresthesia, altered smell or taste, depression or anxiety, gastrointestinal symptoms (diarrhea and abdominal pain), joint or muscle pain, skin rash, and changes in menstrual cycles.^1^ Zhang et al.,^5^ conducted a study with a cohort of 34,605 individuals, using machine learning to analyze over 137 symptoms and conditions in long COVID-19. They identified four reproducible sub-phenotypes: cardiac and renal (1), respiratory, sleep, and anxiety (2), musculoskeletal and nervous system (3), and digestive and respiratory system (4). This classification could assist in the development of future protocols tailored to specific sub-phenotypes, given that varying pathophysiological alterations could be influenced by disparate mechanisms, which may serve as potential therapeutic targets.

The pathophysiology of long COVID-19 remains under investigation, and currently, there is no consensus regarding its underlying mechanisms. However, emerging evidence suggests that the immunomodulatory activities of the nervous system may play a significant role in the development and manifestation of symptoms associated with long COVID-19. An association between long COVID-19 and a dysregulated immune response with chronic inflammation has been demonstrated in several studies. Elevations in pro-inflammatory cytokines such as interleukin-6 (IL-6), tumor necrosis factor-alpha (TNF-α), and interleukin-1 beta (IL-1β) have been observed, indicating a state of sustained inflammation.^6^ Additionally, increased levels of other cytokines, including interleukin-8 (IL-8) and interleukin-10 (IL-10), have been reported, suggesting a complex interplay between pro-inflammatory and anti-inflammatory processes.^6^ C-reactive protein (CRP), an acute-phase reactant produced by the liver in response to inflammation, has also been studied in the context of long COVID-19. Elevated CRP levels have been detected in individuals with persistent symptoms, indicating ongoing systemic inflammation.^7^ Moreover, longitudinal studies have shown a correlation between CRP levels and the severity and duration of symptoms in long COVID-19 patients. The specific profile of inflammatory cytokines and CRP in long COVID-19 may provide insights into the underlying mechanisms of this condition. However, it is important to note that the cytokine profile can vary among individuals, suggesting heterogeneity in the immune response to the virus. Additionally, the temporal dynamics of cytokine and CRP levels throughout the course of long COVID-19 require further investigation to understand the persistence of symptoms and potential therapeutic interventions.^6,7,8^

Post-viral sequelae from numerous viruses, including mumps, human immunodeficiency virus, hepatitis C, Epstein-Barr, Coxsackie type B virus, and Lyme, have been reported to manifest symptoms of dysautonomia.^4^ Many elements of dysautonomia are common in prolonged COVID-19 symptoms. Notably, the increase in sympathetic activity observed in a form of dysautonomia known as Postural Orthostatic Tachycardia Syndrome (POTS), which is characterized by symptoms such as palpitations, chest discomfort, breathlessness, tremulousness, perspiration, pallor, nausea, diarrhea, and a feeling of cold extremities. Concurrently, symptoms associated with cerebral hypoperfusion, including lightheadedness, dizziness, near syncope, alterations in vision and hearing, generalized or lower limb weakness, and cognitive issues, commonly referred to as ‘brain fog,’ are frequently observed. Carmona-Torre et al.,^4^ report an incidence of post covid related dysautonomia of 2.5%. Considering the overlapping symptoms in all phenotypes 2.5% may be an underestimate of the affected population. The role of the sympathetic nervous system in the manifestation of many long COVID-19 symptoms seems evident, suggesting that its therapeutic targeting could be a logical progression in treatment strategies.

Historically, the nervous system and the immune system were thought to have separate functions, however, the works of Elenkov^8^ and Tracey^9^ have demonstrated that the autonomic nervous system (ANS) and the immune system are inseparable. Crosstalk between the ANS and the immune system via afferent and efferent pathways determine the intensity of the inflammatory response, immune response, and local processes in the damaged tissues through positive and negative feedback mechanisms between the two systems. Fischer^10^ states that the ANS plays a coordinating function during viral infections such as SARS COVID-19; the consequences of acute hyperinflammation, endothelial dysfunction, microcirculation disturbance, and coagulopathy are not different entities in a viral infection, but an expression of a malfunction of the ANS with sympathetic hyperactivity.^11^

In the context of long COVID-19, an overactive sympathetic system combined with an underactive vagus nerve could disrupt the balance between these systems, possibly allowing unchecked inflammation to persist. This continued inflammation might be a central element in symptoms characteristic of long COVID-19, autonomic dysfunction with the chronic inflammation resulting in dysregulation of the balance between the sympathetic and parasympathetic systems.

Given the current understanding that emphasizes a significant autonomic component in long COVID-19, we chose to focus our intervention on the sympathetic nervous system. We employed a stellate ganglion block, with the expectation that this approach could potentially alleviate the symptoms of long COVID-19.

## Methods

### Study Design

A cohort population that consisted of patients who received a SGB for the treatment of long COVID-19 symptoms in our Colorado clinics between September and December of 2022. Ultrasound guided injection around stellate ganglion.

### Sampling

Patients who responded to our post-procedure follow-up via phone call, in person or telehealth meeting.

### Data Collection

Data was gathered and stored in an Excel spreadsheet. The data points included 18 items from the CDC list of the most prevalent symptoms associated with long COVID-19 syndrome. For each patient, the data indicated the presence of specific symptoms before the stellate ganglion block (SGB) procedure, and whether these symptoms improved or remained unchanged after the SGB.

### Variables and Measurements

Difficulty breathing or shortness of breath, cough, Tiredness/fatigue, chest or stomach pain, joint or muscle pain, tachycardia/palpitations, symptoms that get worse after physical activity, pins and needles feeling, diarrhea, change in smell and taste, fever, dizziness/lightheadedness when standing, difficulty sleeping, rash, mood changes, headache, changes in menstrual cycle, brain fog/confusion, age and gender. Measured as present or not present before SGB and improved or unimproved after SGB.

### Data analysis

SPSS Version 28.0.1.0 paired sample proportions

### Ethical considerations

All patients included in this study signed an informed consent for the procedure and subsequent collection of de-identified data.

### Limitations

Limited sample size and potential bias of only having the patients who completed post procedural follow up included.

## Narrative

The treatment outcomes of forty-one patients 18 male and 23 female with an age range between 18 and 89 who underwent stellate ganglion block (SGB) for long COVID-19 Syndrome were evaluated. The population consisted of patients from the United States who received treatment in our clinics between September and December of 2022 and responded to our post-procedure follow-up calls. The timeframe between the procedure and the data collection was variable depending on patient response times. The evaluation of the patients’ long COVID-19 symptoms was based on the CDC list of the most prevalent symptoms of the condition.

The initial symptoms reported by the 41 patients include: shortness of breath (41%, 10 males, 7 females), cough (24%, 6 males, 4 females), fatigue (85%, 15 males, 20 females), chest pain (24%, 4 males, 6 females), joint/muscle pain (39%, 7 males, 9 females), tachycardia or palpitations (22%, 4 males, 5 females), post-exertional malaise (66%, 14 males, 13 females), pins and needles sensation (22%, 3 males, 6 females), diarrhea (20%, 3 males, 5 females), changes in taste and smell (44%, 10 males, 8 females), fever (2%, 1 female), dizziness (41%, 11 males, 6 females), difficulty sleeping (34%, 7 males, 7 females), rash (5%, 2 females), mood changes (51%, 11 males, 10 females), headache (39%, 8 males, 8 females), changes in menstrual cycle (12%, 5 females), and brain fog (80%, 15 males, 18 females). (Table 1)

**Table 1:** Long COVID-19 Symptoms Reported This table shows the prevalence of long COVID-19 symptoms reported by patients who underwent stellate ganglion block (SGB) treatment for long COVID-19 syndrome.

| Symptom | Total Reports | Percentage (%) | Males | Females |
| --- | --- | --- | --- | --- |
| Shortness of breath | 17 | 41% | 10 | 7 |
| Cough | 10 | 24% | 6 | 4 |
| Fatigue | 35 | 85% | 15 | 20 |
| Chest pain | 10 | 24% | 4 | 6 |
| Joint/muscle pain | 16 | 39% | 7 | 9 |
| Tachycardia/palpitations | 9 | 22% | 4 | 5 |
| Post-exertional malaise | 27 | 66% | 14 | 13 |
| Pins and needles | 9 | 22% | 3 | 6 |
| Diarrhea | 8 | 20% | 3 | 5 |
| Changes in taste/smell | 18 | 44% | 10 | 8 |
| Fever | 1 | 2% | 0 | 1 |
| Dizziness | 17 | 41% | 11 | 6 |
| Difficulty sleeping | 14 | 34% | 7 | 7 |
| Rash | 2 | 5% | 0 | 2 |
| Mood changes | 21 | 51% | 11 | 10 |
| Headache | 16 | 39% | 8 | 8 |
| Changes in menstrual cycle | 5 | 12% | 0 | 5 |
| Brain fog | 33 | 80% | 15 | 18 |

The dataset, as illustrated in Table 2, presents an assessment of the treatment efficacy in mitigating symptoms. The data indicates noteworthy amelioration in symptoms such as cough, fatigue, and joint pain. There are a substantial proportion of individuals reporting relief, with notable variations in improvement rates between males and females for specific symptoms The findings demonstrate that a substantial proportion of patients (86%) reported a reduction in at least some of their symptoms. Out of the 35 individuals who experienced symptom relief, a majority of 25 patients (61% of the total 41 patients) reported relief from all the long COVID-19 symptoms they initially presented with. (Table 2)

**Table 2:** Long COVID-19 Symptom Resolution. This table displays the percentage of patients who reported resolution of specific long COVID-19 symptoms following stellate ganglion block (SGB) treatment. The symptoms included in this table are fatigue, brain fog, loss of taste and smell, shortness of breath, chest pain, joint pain, muscle pain, headache, and palpitations. The percentage of patients reporting resolution of each symptom is displayed on the y-axis. The x-axis displays the specific symptoms evaluated in this study.

| Symptom | Total Reports | All Relief (%) | Males (Relief %) | Females (Relief %) |
| --- | --- | --- | --- | --- |
| Shortness of breath | 17 | 88 | 80 | 100 |
| Cough | 10 | 80 | 80 | 67 |
| Fatigue | 35 | 77 | 67 | 85 |
| Chest pain | 10 | 80 | 50 | 100 |
| Joint pain | 16 | 94 | 86 | 100 |
| Tachycardia/palpitations | 9 | 78 | 50 | 100 |
| Symptoms worsen | 27 | 74 | 64 | 92 |
| Pins and needles | 9 | 100 | 100 | 100 |
| Diarrhea | 8 | 88 | 67 | 100 |
| Changes in taste/smell | 18 | 56 | 40 | 75 |
| Fever | 1 | 100 | - | 100 |
| Dizziness | 17 | 77 | 64 | 100 |
| Difficulty sleeping | 14 | 71 | 57 | 86 |
| Rash | 2 | 100 | - | 100 |
| Mood changes | 21 | 76 | 64 | 90 |
| Headache | 16 | 81 | 63 | 100 |
| Changes in menstrual cycle | 5 | 80 | - | 80 |
| Brain fog | 33 | 79 | 60 | 94 |

Ultrasound-guided stellate ganglion blocks (SGBs) were performed with patients lying in a supine position with the head of the bed slightly elevated and turned away from the side of the procedure. This positioning, combined with ultrasound guidance, reduces the risk of unintended outcomes, such as pneumothorax, vascular puncture, or cervical nerve intrusion. Elevating the head of the bed helps decompress the subclavian vessels and turning the head away from the procedure side shortens the distance between the needle entry point and the SG. The blocks were performed following standard aseptic procedures. The cricothyroid notch, the border of the sternocleidomastoid muscle, and Chassaignac’s tubercle serve as useful anatomical landmarks during the procedure. A skin marker was used to mark the expected placement of the ultrasound prior to application. The ultrasound probe was positioned transversely at the level of the cricothyroid notch, and the structures of concern were identified, trachea, esophagus, thyroid gland, blood vessels, muscles, and nerves. A lateral to medial in-plane approach was utilized to avoid the structures of concern and to facilitate ideal needle tip placement. Located anterolaterally to the longus colli muscle, deep to the prevertebral fascia, and superficial to the fascia investing the longus colli muscle. A skin wheal with 1 ml of 1% lidocaine with sodium bicarbonate 8% was made, followed by the introduction of a 2 inch or 4-inch echogenic needle (size determined by body habitus) with continuous ultrasound guidance to the area of the SG. A mixture of 2ml 2% Chlorprocaine, 2ml 1% Lidocaine, and 1 ml 0.2% Ropivacaine was used as the injectate, and real-time ultrasound monitoring was used to ensure proper deposition around the SG.^12,13,14,15^

This technique did not result in any significant adverse events. Three patients experienced a vagal response, all of which happened immediately after a left-sided block and resolved quickly with reverse Trendelenburg positioning. Transient hoarseness occurred in four patients and resolved upon dissipation of the local anesthetic, which usually occurred within an hour.

Patients who live locally received a right sided SGB unless their symptoms were purely cardiac in nature. Patients who traveled from out of state, first received a right stellate ganglion block, followed by a left-sided block once the Horner’s syndrome (drooping eyelid, pupil constriction, and absence of sweating) from previous block had resolved.

The resolution of long COVID-19 Symptoms following stellate ganglion block (SGB) was found to occur rapidly, with most patients reporting a noticeable improvement in symptoms within 15 minutes of the block. The resolution of symptoms that could not be clearly evaluated immediately after the SGB (brain fog, fatigue) were observed to occur at different time points, with some patients reporting a steady improvement over one to two weeks, while others reported that their symptoms resolved within a day. However, it should be noted that the current study did not specifically evaluate the time-based resolution of symptoms.

The symptoms most frequently reported by our patients were fatigue, observed in 85% of cases, and brain fog, present in 80% of individuals. These symptoms showed the highest response to treatment, with 77% of patients experiencing fatigue reporting significant relief, and 80% of those suffering from brain fog reporting relief. In our patient group, the loss of taste and smell proved to be the most challenging symptom to treat, with only 56% of patients reporting improvement. Although, we were able to obtain a higher response rate for anosmia with the addition of a trigeminal nerve block.

There was a marked statistical significance across all symptoms in response to stellate ganglion block treatment, with p-values below 0.001 for most symptoms, demonstrating the broad effect of the treatment. The exception was “changes in menstruation”, for which the significance was still noteworthy with a p-value less than 0.05, in both one-sided and two-sided tests. This suggests a comprehensive impact of the stellate ganglion block treatment across a range of symptoms. (Table 3) A gender disparity was observed in both the initial presentation of symptoms and symptom resolution. Women reported a higher frequency of symptoms related to long COVID-19 however, they also demonstrated a higher rate of symptom resolution.

**Table 3:** Tests of Significance for each variable.

| Paired-Samples Proportions Tests |  |  |  |  |  |  |
| --- | --- | --- | --- | --- | --- | --- |
|  | Test Type | Difference in Proportions | Asymptotic Standard Error | Z | Significance |  |
|  |  |  |  |  | One-Sided p | Two-Sided p |
| Pair 1: Difficulty breathing or shortness of breath – Difficulty breathing or shortness of breath Post Treatment | McNemar | .366 | .075 | 3.873 | <.001 | <.001 |
| Pair 2: Cough – Cough Post Treatment | McNemar | .195 | .062 | 2.828 | .002 | .005 |
| Pair 3: Fatigue or tiredness – Fatigue or tiredness Post Treatment | McNemar | .659 | .074 | 5.196 | <.001 | <.001 |
| Pair 4: Chest or stomach pain – Chest or stomach pain Post Treatment | McNemar | .195 | .062 | 2.828 | .002 | .005 |
| Pair 5: Joint or muscle pain – Joint or muscle pain Post Treatment | McNemar | .366 | .075 | 3.873 | <.001 | <.001 |
| Pair 6: Tachycardia or palpitations – Tachycardia or palpitations Post Treatment | McNemar | .171 | .059 | 2.646 | .004 | .008 |
| Pair 7: Symptoms that get worse after physical or mental activities – Symptoms that get worse after physical or mental activities Post Treatment | McNemar | .512 | .078 | 4.583 | <.001 | <.001 |
| Pair 8: Pins and needles feeling – Pins and needles feeling Post Treatment | McNemar | -.780 | .065 | -5.657 | <.001 | <.001 |
| Pair 9: Diarrhea – Diarrhea Post Treatment | McNemar | .171 | .059 | 2.646 | .004 | .008 |
| Pair 10: Changes in taste or smell – Changes in taste or smell Post Treatment | McNemar | .244 | .067 | 3.162 | <.001 | .002 |
| Pair 11: Fevers – Fevers Post Treatment | McNemar | -.976 | .024 | -6.325 | <.001 | <.001 |
| Pair 12: Dizziness of lightheadedness when standing – Dizziness of lightheadedness when standing Post Treatment | McNemar | .317 | .073 | 3.606 | <.001 | <.001 |
| Pair 13: Difficulty sleeping – Difficulty sleeping Post Treatment | McNemar | .244 | .067 | 3.162 | <.001 | .002 |
| Pair 14: Rash – Rash Post Treatment | McNemar | -.951 | .034 | -6.245 | <.001 | <.001 |
| Pair 15: Mood changes – Mood changes Post Treatment | McNemar | .390 | .076 | 4.000 | <.001 | <.001 |
| Pair 16: Headache – Headache Post Treatment | McNemar | .317 | .073 | 3.606 | <.001 | <.001 |
| Pair 17: Brain fog or confusion – Brain fog or confusion Post Treatment | McNemar | .634 | .075 | 5.099 | <.001 | <.001 |
| Pair 18: Changes in menstruation – Changes in menstruation Post Treatment | McNemar | .098 | .046 | 2.000 | .023 | .046 |

## Discussion

The precise mechanism through which SGB ameliorates long COVID-19 symptoms is not fully understood. Fisher et. al.^10^ theorizes that the immune and inflammatory responses during the acute phase of long COVID-19 and the ensuing post-acute symptoms are due to a malfunction in the Autonomic Nervous System (ANS), particularly sympathetic hyperactivity. They argue that SGB, by modulating the ANS, can disrupt overactive processes within nerve-immune-inflammation feedback loops, enabling their self-regulation. Furthermore, SGB’s impact goes beyond the ANS, influencing downstream regulatory actions including immune and inflammatory system control, cytokine production, endothelial function, and microcirculatory function regulation.^10^ SGB has been utilized for a broad spectrum of conditions, including pain syndromes and disorders of the immune and endocrine systems. Lipov et al.,^16^ suggested that the therapeutic action of SGB might primarily involve sympathetic block-mediated peripheral vasodilation. However, given the diversity of SGB applications, they acknowledge that the mechanism might be more multifaceted. Deng et al.,^17^ demonstrated multiple changes in the neuroendocrine system of rats that received an SGB prior to surgery. The changes included reductions in neuronal loss, decreased microglial activation, as well as reductions in inflammatory markers TNF-α, IL-1β, and IL-6. Others have hypothesized that SGB results in changes in voltage-gated sodium channels of peripheral nerves and the central response by spinal feedback loops, thus decreasing symptoms.^4^ Neuronal growth factor is also suspected to play a role in the effects of a SGB.^18^ Multiple studies and publications have shown that SGB has the following effects in regulating the immune system and hyperinflammation:

Reduction of natural killer cell activity.^19, 20^
Reduction of inflammatory cytokines IL-1, IL-4, IL-6, IL-8, TNF-α.^21, 22, 23^
Increase of anti-inflammatory cytokine IL-10 and of CGRP.^21, 22, 23^
Regulation of the endothelial dysfunction.^24^
Regulation of microcirculation and coagulopathy.^20, 25, 26^
Reduction of (neurogenic) pulmonary edema.^15, 24, 27^
Reduction of pulmonary arterial hypertension.^28^
Reduction of pathological positive feedback loop.^10, 29, 30, 31^

The heightened sympathetic activity associated with long COVID-19 may lead to vasoconstriction, which can decrease cerebral blood flow (CBF). The application of SGB could induce vasodilation, increasing CBF and potentially ameliorating symptoms. While some research suggests that SGB could enhance CBF by curtailing sympathetic activity, other studies propose that its impact could be negligible in healthy individuals due to the brain’s inherent autoregulatory mechanisms.^32^ Nonetheless, the exact effect of SGB on CBF remains an active area of investigation.

The reversal of anosmia (loss of smell) and dysgeusia (altered taste) post-Stellate Ganglion Block (SGB) aligns with the restoration of parasympathetic function due to sympathetic blockade of the Stellate Ganglion (SG), the conduit for all sympathetic innervation of the head and neck. It is plausible to infer that the sympathetic hyperreactivity, also triggers anosmia and dysgeusia by interrupting sympathetic outflow to cranial nerves, specifically CN I (Olfactory), CN VII (Facial), and CN IX (Glossopharyngeal).^14^ Blocking the SG, and subsequently the superior and middle cervical ganglion, impacts post-ganglionic nerves that connect to the internal carotid nerve, external carotid nerve, nerve to the pharyngeal plexus, nerves to cranial nerves II, III, IV, VI and IX, and gray rami communicantes.^33^

The timing for employing SGB in managing long-COVID syndrome may be important, as the degree of neuroadaptation that has occurred may decrease the effectiveness of the SGB. Our clinical observations indicate that delayed SGB, such as in individuals with long standing analogous post-viral ailments (Lyme Disease, Chronic Fatigue Syndrome, Myalgic Encephalopathy) exhibited notably diminished responsiveness to SGB as compared to long COVID-19 patients. Achieving only a 10% - 20% reduction in their symptoms with each injection.

In our practice, SGB has proven beneficial in treating long COVID-19 symptoms, and we urge its prompt consideration to avert possible diminished effectiveness due to time-sensitive neuroadaptive changes.

This study’s results, based on responses from 41 of 60 treated patients, may not wholly represent all patient experiences. Those with less positive outcomes might have opted out of participating.

The choice of local anesthetic could influence SGB efficacy. We used a mix of Chloroprocaine, Lidocaine, and Ropivacaine. Lidocaine is known for its immunomodulatory properties, while others have not been tested in this context. Further research might help determine the most effective local anesthetic and the underlying therapeutic mechanisms of SGB in long COVID-19 patients.

The use of ultrasound guidance during SGB injections is advocated as it helps to avoid critical structures and reduce the risk of needle placement or injection into unintended structures. In our practice, no serious adverse events were reported from SGB injections performed under ultrasound guidance.

In our clinical practice, right-sided stellate ganglion blocks (SGBs) were the most frequently performed procedure. Bilateral SGBs were reserved for patients who were coming from out-of-state or experiencing predominately dysautonomia related to long COVID-19 syndrome. The physiological effects of SGBs can vary depending on the side of injection. Right-sided SGBs have been shown to result in greater sympathetic blockade than left-sided SGBs, leading to a greater decrease in heart rate and blood pressure, likely due to the right stellate ganglion’s dominance and more direct connection to the heart via the cardiac plexus. ^9, 18, 34, 35,^ In contrast, left-sided SGBs produce a greater degree of parasympathetic effect than right-sided SGBs, leading to a greater increase in heart rate and cardiac output, likely due to the left vagus nerve’s stronger influence on the heart and its direct connection to the sinoatrial node.^9, 18, 34, 35, 36^ While no data were collected to analyze the relationship between laterality and patient response, we believe it is important to investigate the optimal laterality for administering SGBs based on specific symptoms of long COVID-19.

Six patients failed to experience symptom relief, and further evaluation revealed that three had rare disorders that were contributing to their symptoms. It is noteworthy that these diagnoses were uncommon, leading us to question if there may be undiagnosed underlying medical conditions in some of the remaining patients who were unresponsive to treatment.

## Conclusion

The results of our cohort study indicate that SGB is a promising treatment option for patients suffering from long COVID-19 syndrome. The rapid resolution of symptoms and high success rate of treatment make SGB a compelling treatment option for long COVID-19 patients. However, it should be noted that our findings are limited to a specific subset of patients, and further research is needed to confirm these results. The use of ultrasound guidance and the specific anesthetic used may also play a role in the efficacy of SGB, and further studies in these areas are warranted.

While further double-blind placebo-controlled testing is necessary, the highly favorable patient response to this treatment suggests that it should not be withheld by healthcare providers. Despite the need for additional studies, we believe that this safe and effective procedure should be made broadly available to patients suffering from long COVID-19.

## Data Availability

All data produced in the present study are available upon reasonable request to the authors

---

**Figure 1:**
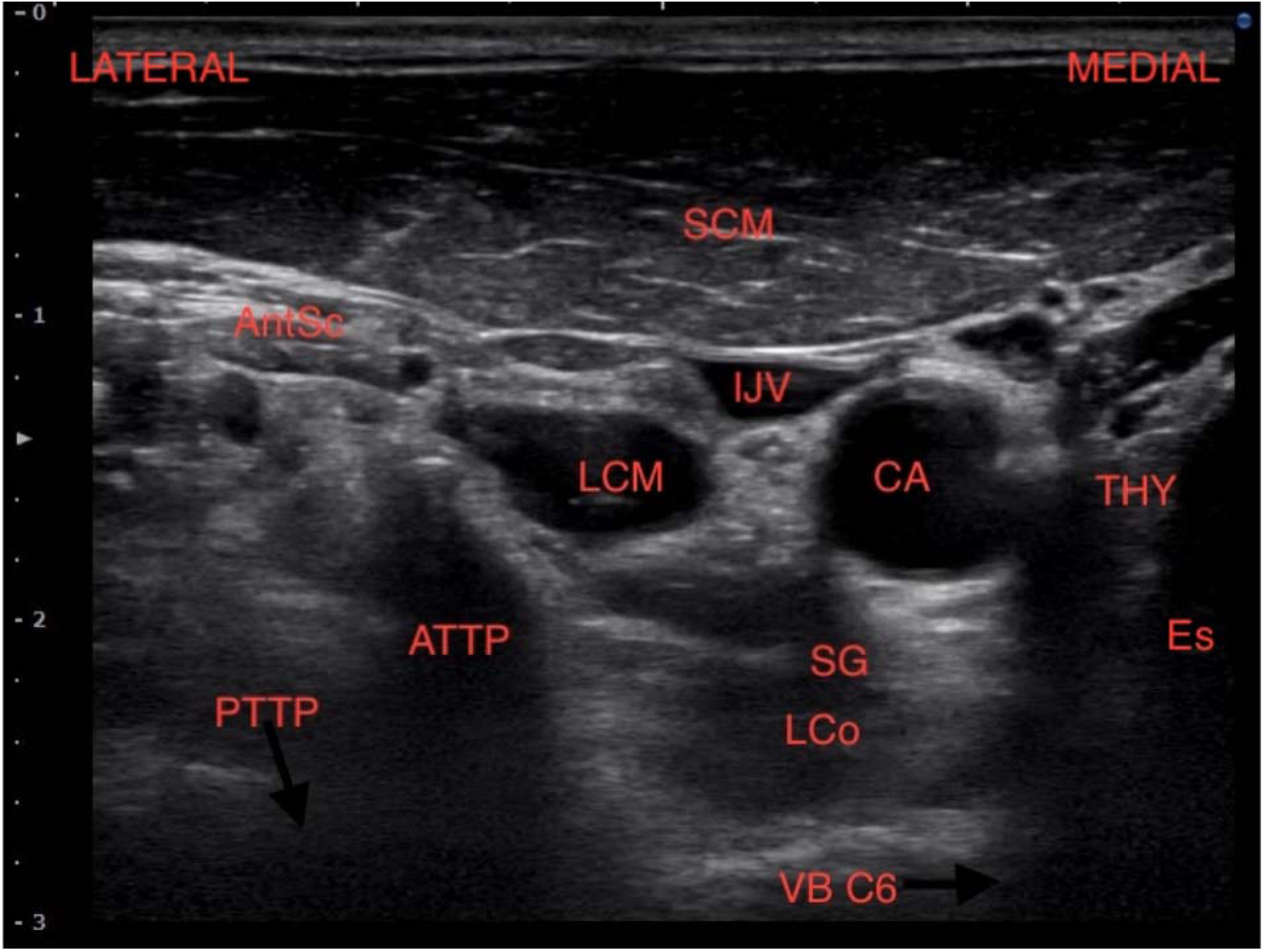
Is an ultrasound image of structures of interest for a SGB. Depicting: C-6 Nerve Root (labeled "C6 NR") Anterior Transverse Process (labeled "ATTP") Posterior Transverse Process (labeled "PTTP") Longus Colli Muscle (labeled "LC") Inferior Thyroid Artery (labeled "ITA") Internal Jugular Vein (labeled "IJV") Carotid Artery (labeled "CA") Thyroid (labeled "Thy")

## Author Contributions

^1^ Lisa Pearson; Principal author, interventionalist, design implementation, data collection, assimilation, and statistical calculation

^2^Alfred Maina; Co-author, interventionalist, data collection

^3^ Taylor Compratt, Data collection

^4^ Sherri Harden, Data collection, assimilation, and statistical calculation

^5^ Abbey Aaroe; Data collection tool development, data collection

^6^ Leah Thompson; Patient Evaluations, Data collection

## Acknowledgements

Thank you for the data collection and article proofreading of:

Whitney Copas, BSN, RN

Victoria King, MD, MHA

John Maye, PhD, CRNA

